# Wastewater-Based Epidemiology and Whole-Genome Sequencing for Community-Level Surveillance of SARS-CoV-2 in Selected Urban Communities of Davao City, Philippines: A Pilot Study

**DOI:** 10.1101/2021.08.27.21262450

**Authors:** Maria Catherine B. Otero, Lyre Anni E. Murao, Mary Antoinette G. Limen, Paul Lorenzo A. Gaite, Michael G. Bacus, Joan T. Acaso, Kahlil Corazo, Ineke E. Knot, Homer Sajonia, Francis L. de los Reyes, Caroline Marie B. Jaraula, Emmanuel S. Baja, Dann Marie N. Del Mundo

## Abstract

**Background:** Over 50 countries have used Wastewater-Based Epidemiology (WBE) and Whole-Genome Sequencing (WGS) of SARS-CoV-2 for monitoring COVID-19 cases. COVID-19 surveillance in the Philippines relies on clinical monitoring and contact tracing, with both having limited use in early detection or prediction of community outbreaks. Thus, complementary public health surveillance methods that can provide community-level infection data faster and using lesser resources must be explored.

**Objectives:** This study piloted and assessed WBE and WGS as approaches for COVID-19 surveillance in low-resource and low-sanitation communities in Davao City, Philippines.

**Methods:** Weekly wastewater samples were collected from six barangay community sewer pipes or creeks from November to December 2020. Samples were concentrated using a PEG-NaCl precipitation method and analyzed by RT-PCR to detect the SARS-CoV-2 N, RdRP, and E genes. In addition, SARS-CoV-2 RNA-positive samples were subjected to WGS for genomic mutation surveillance. Public data from clinical surveillance were also reviewed to interpret WBE data.

**Results:** Twenty-two of the 24 samples (91.7%) obtained from the six barangays tested positive for SARS-CoV-2 RNA. The cycle threshold (Ct) values were correlated with RNA concentration and attack rate. Thirty-two SARS-CoV-2 mutations were detected in WGS, including novel non-synonymous mutations or indels in seven SARS-CoV-2 genes and ten mutations previously reported in the Philippines.

**Discussion:** SARS-CoV-2 RNA was detected in community wastewater from the six barangays of Davao City, even when the barangays were classified as having a low risk of COVID-19 transmission and no new cases were reported. Despite the fragmented genome sequences analyzed, our genomic surveillance in wastewater confirmed the presence of previously reported mutations while identifying mutations not yet registered in clinical surveillance. The local context of a community must be considered when planning to adopt WBE and WGS as complementary COVID-19 surveillance methodologies, especially in low-sanitation and low-resource settings.

## INTRODUCTION

Following the World Health Organization declaration of a global pandemic in March 2020, public health surveillance systems at the national and local levels were immediately instituted to reduce the transmission of COVID-19. Surveillance aims to rapidly detect, isolate, and manage cases and their contacts, guide control measures, and monitor epidemiologic trends and evolution of SARS-CoV-2 (World Health Organization, 2020). In the Philippines, COVID-19 surveillance relies heavily on RT-PCR-based clinical diagnostics in symptomatic individuals and their contacts. However, clinical monitoring and contact tracing have limited use in the early detection or prediction of community outbreaks and can be logistically demanding when applied to a large population (Sims and Kasprzyk-Horden, 2020; Nsubuga et al., 2006). When interventions aim to curb transmission from patients with no symptoms or with a mild infection, complementary public health surveillance methods that can provide faster results of community-level infection data using lesser resources must be explored.

SARS-CoV-2 is shed in the feces of symptomatic and asymptomatic COVID-19 cases (Chen et al., 2020; Park et al., 2020; Schmitz et al., 2021 PREPRINT). Over 50 countries have utilized sewage wastewater monitoring for COVID-19, whether in national or regional scale, or in localized sites (Naughton et al., 2021 PREPRINT). Studies from the Netherlands and the USA showed that wastewater-based epidemiology (WBE), or wastewater-based surveillance, can detect and predict hotspots of COVID-19 infections earlier than clinical surveillance (Wu et al., 2020; Wurtzer et al., 2020). Larsen and Wigginton (2020) further projected that WBE could give a 7-day lead over COVID-19 case reporting in symptomatic or clinical surveillance. Thus, WBE can serve as an early warning tool that can help control the spread of COVID-19, especially in areas where diagnostic resources are limited (Bivins et al., 2020). WBE can also be utilized in assessing control strategies against COVID-19 at the community level. SARS-CoV-2 viral titers in WBE declined in response to social isolation and lockdowns in Montana, USA (Nemudryi et al., 2020), Paris, France (Wurtzer et al., 2020 PREPRINT), and Rio de Janeiro, Brazil (Prado et al., 2021). In Brazil (Prado et al., 2021) and Singapore (Wong et al., 2021), WBE data from samples from sewer pipes were used to determine where public health interventions should be intensified. Circulating variants and strains of SARS-CoV-2 were identified in wastewater using whole-genome sequencing (WGS), supporting the use of wastewater surveillance for tracing variant importations and circulation (Martin et al., 2020; Crits-Christoph et al., 2021; Napit et al.,2021 PREPRINT).

Davao City is the third most populous Highly Urbanized City (HUC) in the Philippines, with 1.8 million inhabitants. It is considered as the gateway for trade in the Southern Philippines (PSA, 2019). There were 8,808 confirmed cases of COVID-19 in Davao City at the end of 2020, coming from 178 of its 182 *barangays* (or smallest administrative division). Like many cities in the Philippines, Davao City does not have a centralized wastewater management system. As a result, inadequately treated domestic wastewater and sewage from private and communal septic tanks may be released to rivers and creeks along with surface run-off (Guerrero-Latorre et al., 2020; JICA, 2018). COVID-19 WBE in low-sanitation settings in Ecuador (Guerrero-Latorre et al., 2020) and portions of Brazil (Prado et al. 2020) demonstrated how data from WBE could be used in designing public health interventions and monitoring COVID-19 transmission.

This study piloted and assessed WBE and WGS as methodologies for COVID-19 surveillance in low-resource and low-sanitation communities in Davao City, Philippines. It also identified the challenges in conducting wastewater-based surveillance in low-resource and low-sanitation communities.

## METHODS

### Study Site

This descriptive pilot study was implemented in Davao City, a chartered city in the Davao Region, the southern portion of the Philippines (Figure 1). The City Government of Davao City releases weekly COVID-19 transmission risk categories (CRITICAL, HIGH, MODERATE, or LOW) for its 182 *barangays*. These weekly characterizations are based on new and active cases normalized against Weekly or Average Daily Attack Rate (ADAR) and the 2-week Growth Rate (2WGR) for COVID-19 (City Government of Davao, 2021).

**Figure 1.**
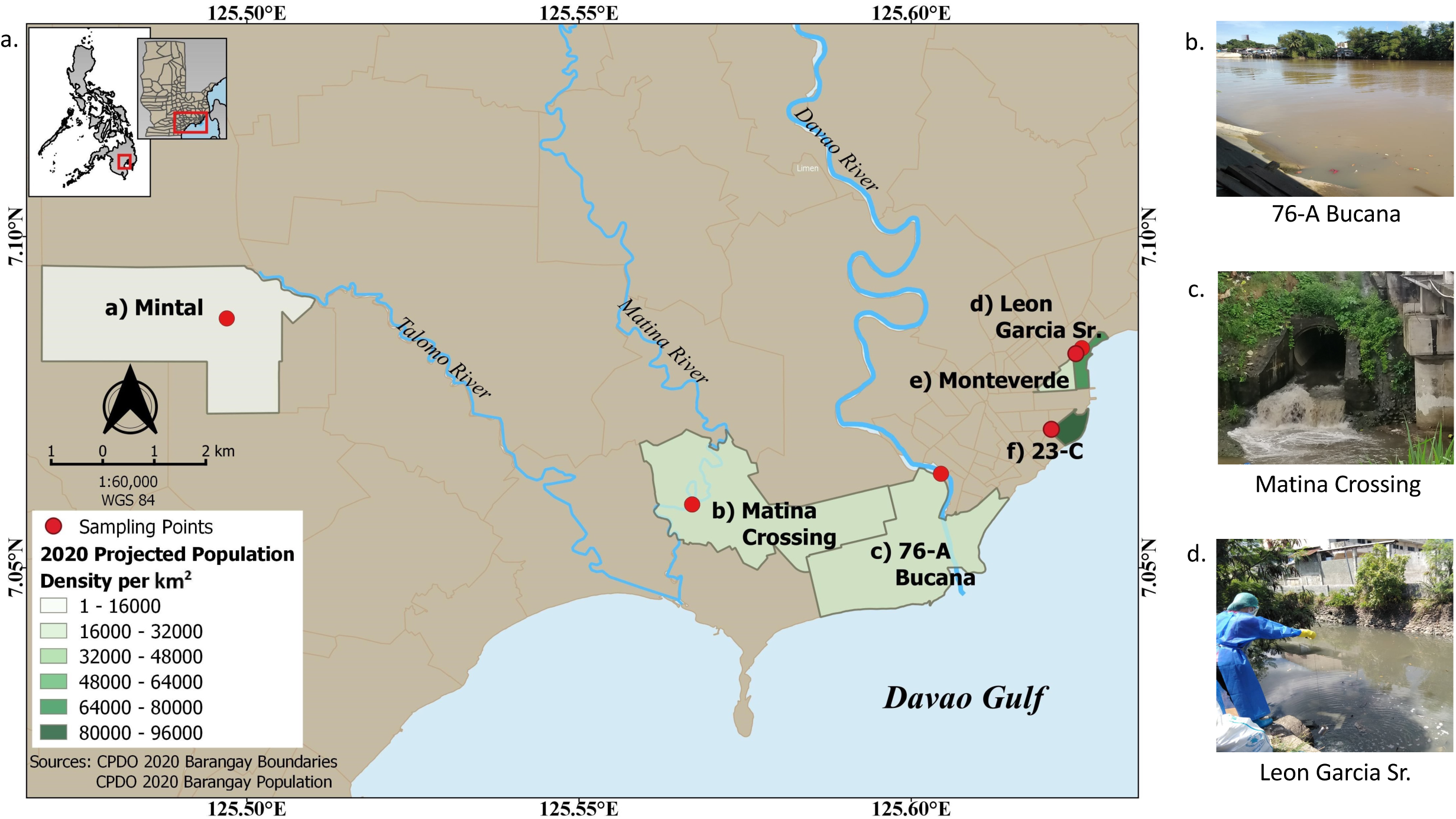
Wastewater outfall and sampling sites in Davao City. a) Location of the six *barangays* and sampling sites with respect to the major draining natural bodies of water. b-d) Different site conditions showing the access points to sewer pipes.

Guided by this weekly COVID-19 Risk Assessment *Barangay* Classification and the expert advice of the City Health Office, six communities with HIGH to MODERATE risk of COVID-19 transmission were identified for sampling (Figure 1). One wastewater sampling site within each *barangay* was determined with the help of the *barangay* personnel. Accessibility and presence of a continuous flow of wastewater were considered in choosing the site. In addition, two tertiary-level hospitals were also included as reference sites. The study protocol was reviewed and approved by an independent Research Ethics Board (BMH-REC Code # 2020-08-02).

### Review of Secondary Data

The communities under each *barangay*, the estimated population as of 2020, and their land area were obtained from the involved *barangays* and the City Planning and Development Office (CPDO) of the City Government of Davao. In addition, clinical surveillance data for COVID-19 was constructed from the publicly available daily list of new cases released by the Department of Health Davao Region (2021). Moreover, new and active cases (defined as new cases in the preceding two weeks) and cumulative incidence per 1,000 persons were computed based on the constructed database.

### Sample collection and processing

Wastewater was collected from the sewer pipes in [1] 23-C, [2] 76-A Bucana, [3] Matina Crossing, and [4] Leon Garcia Sr., and from the creeks in [5] Mintal and [6] Tomas Monteverde between 8 AM to 3 PM once weekly for four weeks from November 8 to December 12, 2020. Weekly sampling was divided into two days, and *barangay* wastewater target sites in close proximity were sampled on the same day. Two hundred fifty milliliters of grab wastewater samples from each site were placed in sterile high-density polyethylene (HDPE) bottles, transported on ice to the College of Science and Mathematics, University of the Philippines Mindanao, and processed in a Biosafety Level 2 laboratory within four hours of collection.

The methods for processing wastewater samples were modified from Wu et al. (2020). Briefly, wastewater samples were sonicated at 20kHz without pulse for two mins. Samples were centrifuged at 4,700xg for 30 mins without brake to remove large particles. The supernatant was decanted and then passed through a 0.22 µm polyethersulfone (PES) syringe filter. Polyethylene glycol (PEG 8000) and NaCl were then added to the filtrate to a final concentration of 8% w/v PEG and 0.3M NaCl. After vigorous agitation, the tubes were kept at 4°C for 20 hours and then centrifuged at 10,000xg for 90 mins until a visible pellet formed. The viral pellet was then reconstituted with 100uL DNA/RNA Shield (Zymo Research, USA) and stored at -20°C until RNA extraction.

### RNA Extraction and RT-PCR analysis

Viral RNA was extracted from 100μL resuspended viral pellets using the MagMax Viral/Pathogen Nucleic Acid Isolation Kit (Thermo Fisher Scientific Inc., USA) in a Kingfisher Duo Prime Purification System (Thermo Fisher Scientific Inc., USA). RT-PCR detection of SARS-CoV-2 was performed using the Allplex™ 2019-nCoV Assay (Seegene Inc., South Korea), which targets the RdRP, N, and E genes. Five microliters each of the 2019-nCoV MOM, RNase-free water, and 5x real-time one-step buffer, 2μL of real-time one-step enzyme, and 8μL of RNA sample or controls (non-template control and positive control) were added for a total qPCR reaction of 25uL. Thermal cycling was set at 50°C for 20 min for reverse transcription, 95°C for 15 min, and then 45 cycles of 94°C for 15s, and 58°C for 30s using the Applied Biosystems QuantStudio 5 real-time PCR machine (Thermo Fisher Scientific Inc., USA). The cut-off cycle threshold (Ct) for Allplex™ 2019-nCoV Assay was set at 40 cycles. Samples were considered POSITIVE for SARS-CoV-2 RNA when all three targets were amplified, OR when either N or RdRP genes, OR both N and RdRP genes were amplified with Ct below or equal to 40, as recommended by the manufacturers in the analysis and interpretation of the Allplex™ 2019-nCoV Assay. However, if only the E gene was amplified, the sample was considered NEGATIVE for SARS-CoV-2 RNA. Linear regression analysis using the IBM SPSS Software (version 21) was used to assess if the Ct of the E, N, and RdRP genes were correlated with the RNA concentration and the computed weekly Attack Rate.

### Whole Genome Sequencing

Eleven (11) wastewater samples that were positive for all targets (RdRP, N, and E genes) and with Ct values ranging from 29 to 39 were chosen for WGS (Supplemental Material, Sample details). SARS-CoV2 amplicon enrichment was performed using the NEBNext SARS-CoV2 companion kit (New England BioLabs, USA) using the IDT 2019-nCoV v3 panel. PCR products for each sample, including a negative control, were diluted to 50µL using 9.5µL of cDNA from each of the two separate primer pool reactions and 31µL nuclease-free water. A total of 5µL PCR dilution per sample was subjected to downstream processing consisting of a DNA end-repair step, barcode ligation, adapter ligation, and library clean-up following the published ARTIC nCoV-2019 sequencing protocol (LoCost) v3 (Quick, 2020). Sequencing using a FLO-MIN106 flow cell on the MinION Mk1B device (Oxford Nanopore Technologies, United Kingdom) was allowed to run for approximately 24 hours.

### Bioinformatic Analysis

Bioinformatic analysis was performed following the nCoV-2019 novel coronavirus bioinformatics protocol by the ARTIC Network (Loman et al., 2020), with several modifications made according to the actual experimental conditions. Briefly, MinKNOW software was used to generate raw sequencing read files in FAST5 file format. The generated FAST5 files were subsequently base called and converted to FASTQ files using the guppy_basecaller software (v.9.4.1) set to high-accuracy mode basecalling. The basecalled reads were then demultiplexed into their corresponding barcodes with the guppy_barcoder software. All barcoded reads were subjected to read filtering through the ARTIC guppyplex tool, retaining reads that are 400 (approximate amplicon size) to 600 bases in length. The remaining reads were then subjected to the medaka pipeline (set to v.9.4.1 as this is the version of the guppy_basecaller used) set to use the ARTIC Protocol V3 primer set, which generated the assembled consensus sequences and performed variant calling. All consensus sequences were submitted to the CoV-GLUE online server bioinformatics tool for corresponding lineage assignment and identification of viral mutations using the SARS-CoV-2 isolate (Wuhan-Hu-1) complete genome (GenBank Accession ID: MN908947.3) as the reference sequence (Singer *et al*., 2020). As the genome assemblies in this study were incomplete and highly fragmented due to the presence of many N base calls and hence sequence gaps, the threshold for variant calling was performed with a total of 30x coverage (Izquierdo-Lara *et al*., 2021) on a specific base across all assembled consensus sequences to ensure high-quality variant calls.

## RESULTS

### Site characteristics

The largest *barangay* by land area is Mintal (7.68 sq. km), and the smallest is 23-C (0.2 sq. km). However, the population density was highest in 23-C with 94,111 persons/sq km, and lowest in Mintal with 1,929 persons/sq km (Table 1). Community wastewaters included in this study converged in barangay sewer lines that drained either to a river (Talomo River, Matina River, and Davao River) or to the Davao Gulf (Figure 1).

**Table 1.** Barangay and sampling site characteristics.

| Barangay | 2020 Est.<br>Population | Land<br>Area<br>(sq. km.) | Population<br>Density<br>(persons/s<br>q. km.) | Source of<br>wastewater | Water body<br>receiving<br>wastewater | New<br>COVID-19<br>cases<sup>1</sup> | Active<br>COVID-19<br>cases<sup>1</sup> |
| --- | --- | --- | --- | --- | --- | --- | --- |
| 23-C | 18,474 | 0.20 | 94,111.05 | Sewer pipe | Davao Gulf | 17 | 23 |
| Leon Garcia | 15,296 | 0.22 | 68,224.80 | Sewer pipe | Davao Gulf | 44 | 59 |
| Monteverde | 6,404 | 0.21 | 31,042.17 | Creek | Davao Gulf | 20 | 26 |
| 76-A<br>Bucana | 94,074 | 4.02 | 23,409.06 | Sewer pipe | Davao River | 198 | 264 |
| Matina<br>Crossing | 36,342 | 5.29 | 6,866.57 | Sewer pipe | Matina<br>River | 149 | 196 |
| Mintal | 14,820 | 7.68 | 1,929.16 | Creek | Talomo<br>River | 22 | 31 |
| Entire<br>Davao City | 1,816,987 | 2,440.00 | 744.67 | - | - | 3,120 | 4,379 |

<sup>1</sup> From Nov 8 to Dec 12, 2020

Except for Leon Garcia Sr., all barangays were at a high-risk level of COVID-19 transmission at least once during the study period (Figure 2). In addition, the following barangay, 23-C, Leon Garcia Sr., Tomas Monteverde, and Mintal were categorized as low-risk barangays during one of the four weeks of wastewater testing (Figure 2A, C, E, and F). None of the barangays were categorized as very high or critical risk during the entire wastewater sampling. Moreover, no risk assessment was declared for the week of Nov 15 to 21, 2020.

**Figure 2.**
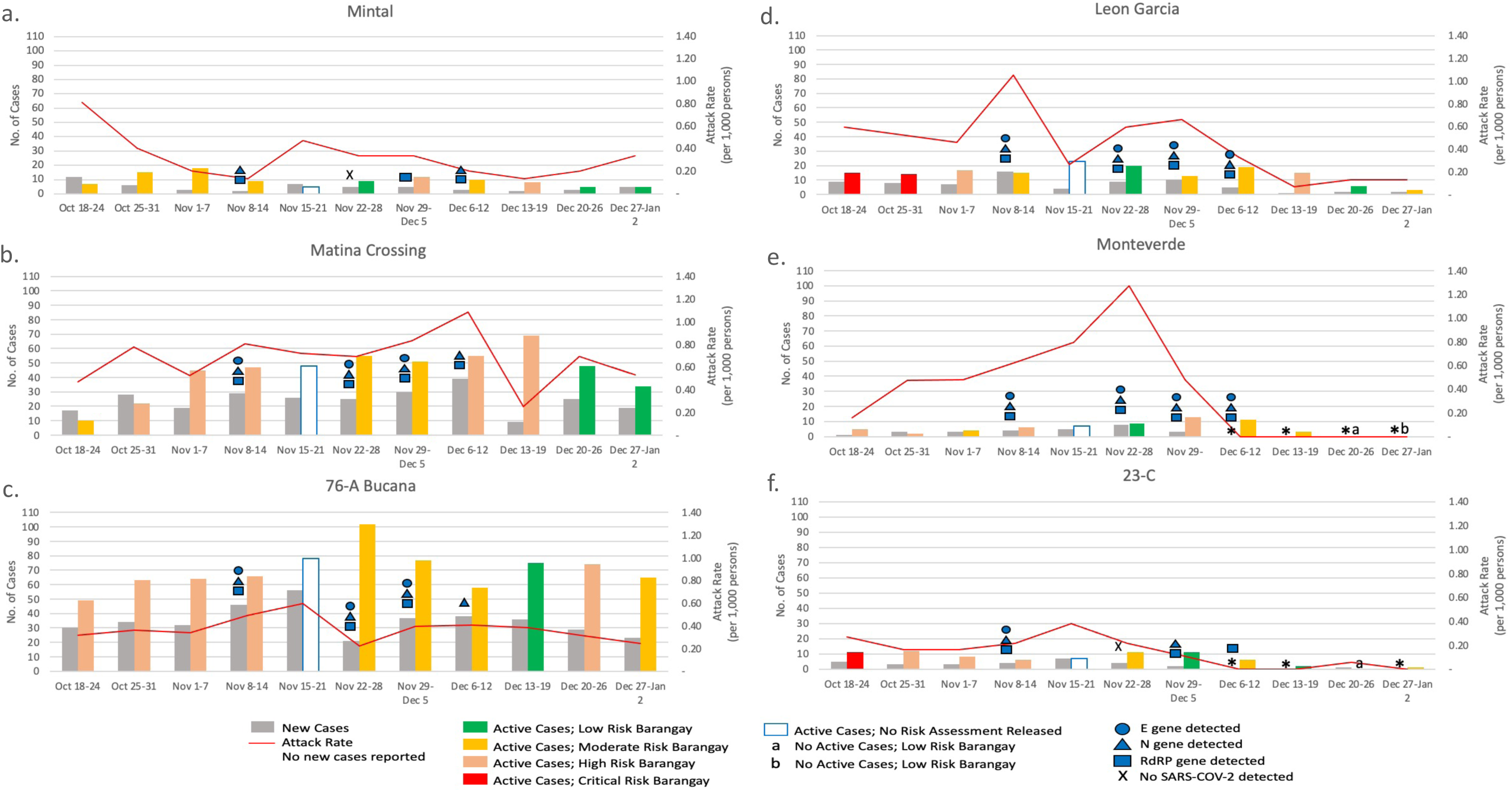
COVID-19 status and SARS-CoV-2 wastewater surveillance in six barangays of Davao City, Philippines.

Four hundred fifty new and 599 active cases were reported in the six *barangays* during the wastewater testing period (Table 1). These cases cover only 14% of the new COVID-19 cases and 14% of the active COVID-19 cases reported in the entire Davao City for the same period. Over 44% of the new cases in the six *barangays* occurred in 76-A Bucana. In addition, the weekly number of new and active cases was consistently higher for 76-A Bucana and Matina Crossing relative to the other barangays (Table 1 and Figure 2). The weekly attack rates ranged from 5 to 127 new cases per 100,000 persons, with the highest attack rate recorded in Tomas Monteverde on November 22 to 28 (Figure 2).

### SARS-CoV-2 Detection in Wastewater

Twenty-two out of the 24 (91.7%) samples obtained from the six barangays over the four-week course of the study tested positive for SARS-CoV-2, with RNA concentrations ranging from 7.8 to 40.2 ng/uL (Supplemental Material, Sample details) and Ct values ranging from 29.41 to 39.73 (Table 2). Moreover, the detection rate for the individual genes was highest for RdRP at 87.5%, followed by N at 83.3% and E at 62.5%. However, no SARS-CoV-2 genes were detected in 23-C and Mintal samples with low recovered RNA levels of 7.8 ng/uL RNA and 10.6 ng/uL RNA, respectively, during the week of November 22 to 28, 2020. In addition, SARS-CoV-2 RNA was also detected in all untreated wastewater samples from the two reference tertiary level hospitals in Davao City, with Ct values ranging from 30.95 to 39.01 (Table 2).

**Table 2.** SARS-CoV-2 detection in six barangays of Davao City, Philippines on November 8-Dec 12, 2020.

| Gene Marker | Communities<sup>1</sup> |  | Hospitals<sup>2</sup> |  |
| --- | --- | --- | --- | --- |
|  | Positive (%) | Ct range<sup>3</sup> | Positive (%) | Ct range<sup>3</sup> |
| **E** | 15/24 (62.5%) | 29.95 - 39.73 | 5/8 (62.5%) | 33.03 - 37.64 |
| **N** | 20/24 (83.3%) | 29.41 - 38.74 | 8/8 (100%) | 30.95 - 37.48 |
| **RdRP** | 21/24 (87.5%) | 31.26 - 38.89 | 6/8 (75%) | 31.67 - 39.01 |
| **Overall<sup>4</sup>** | 22/24 (91.7%) | 29.41 - 39.73 | 8/8 (100%) | 30.95 - 39.01 |

<sup>1</sup> Total of 24 samplings
<sup>2</sup> Total of eight samplings
<sup>3</sup> Ct values of positive samples
<sup>4</sup> Based on the criteria of Allplex SARS-CoV-2 Assay

### Epidemiologic Assessment of SARS-CoV-2 Wastewater Surveillance

SARS-COV-2 RNA was detected in all sites in at least three out of four sampling points between November and December 2020, regardless of the *Barangay* Risk Category and even for *barangays* with a weekly attack rate as low as one new case per 10,000 population (Figure 2). Notably, SARS-CoV-2 RNA was detected in 23-C and Monteverde for the week of December 5-12, even when no new cases were reported for the corresponding week (Figure 2A and E). However, SARS-CoV-2 RNA was not detected even if new cases were reported in 23-C for November 22-28 with a weekly attack rate of 22 per 100,000 persons and in Mintal for November 15-21 with a weekly attack rate of 47 per 100,000 persons (Figure 2A and F). Furthermore, the Ct for the E and N genes was negatively correlated with RNA concentration and attack rate (p-values < 0.05). However, the Ct value RdRP gene was only associated with the changes in the attack rate (p-value 0.04, Supplemental Material, regression analyses per gene target).

### SARS-CoV-2 Sequencing in Wastewater

Whole-genome sequence data from SARS-CoV-2-positive wastewater samples showed that genome coverage varied widely among samples. The November 9 sample from Matina Crossing had ∼50% genome fraction coverage. All other samples had a range of 1.16% - 20.37% genome fraction coverage, indicating incomplete and highly fragmented assembled genomes. CoV-GLUE analyses of the assembled consensus genome sequences assigned these sequences to differing lineages, such as B.1, B.1.1, and B.6 (Supplemental Material, genome assembly and analyses). However, the likelihood weight ratio values for these lineage assignments ranged from 59.375 - 100, with five samples having values below 75, most likely due to the highly fragmented nature of the assembled genomes.

CoV-GLUE detected a total of 32 mutations among all assembled consensus sequences, nine of which were synonymous, 17 were non-synonymous, five were indels, and one was located in a non-coding region (Table 3). Most of the mutations were detected in only one sample, a few in two samples, and one in six samples. Ten of these mutations have already been previously reported, but novel non-synonymous mutations or indels in the coding regions of nsp3, nsp6, RdRp, ORF3, ORF6, ORF7A, and ORF8 were also noted. Among the known mutations in the spike gene for SARS-CoV-2 Variants of Concern (VOCs) and the Philippine P3 variant, only D614G was detected (Table 3 and Supplemental Material, gene mutations). The S94F, 141-143del, P681H/R, K417N/T, E1092K, and H1101Y mutations were absent among samples whose sequence data covered these regions. The other mutations were not covered in the sequence data.

**Table 3.** Information on variant calling and corresponding mutations of SARS-CoV-2 detected from wastewater in Davao CIty, Philippines on Nov-Dec 2020.

| Base Position in Reference Sequence | Reference Base | Alternate Base | Type of Mutation | Resulting Mutation | Affected Gene/ Protein/ Region | Number of Sample(s) Represented | Total Number of Observations | Previously Reported? |
| --- | --- | --- | --- | --- | --- | --- | --- | --- |
| 1190 | C | T | Non-synonymous | P129S (P309S) | nsp2 (ORF1a) | 1 | 155 | Yes, Sengupta *et al.* (2020) |
| 5469 | T | C | Non-synonymous | L917S (L1735S) | nsp3 (ORF1a) | 1 | 31 | No |
| 6312 | C | A | Non-synonymous | T1198K (T2016K) | nsp3 (ORF1a) | 1 | 84 | Yes, Tablizo *et al.* (2020) |
| 8299 | C | T | Synonymous | N1860 (N2678) | nsp3 (ORF1a) | 1 | 155 | No |
| 11082 | TG | T | Deletion | Frameshift | nsp6 (ORF1a) | 2 | 605 | No |
| 11781 | A | G | Non-synonymous | K270R (K3839R) | nsp6 (ORF1a) | 1 | 72 | No |
| 13730 | C | T | Non-synonymous | A97V | RdRP | 2 | 40 | Yes, Tablizo *et al.* (2020) |
| 14168 | C | T | Non-synonymous | P243L | RdRP | 1 | 81 | No |
| 14188 | G | T | Non-synonymous | A250S | RdRP | 1 | 67 | No |
| 14786 | C | T | Non-synonymous | A449V | RdRP | 1 | 134 | Yes, Wang *et al.* (2020) |
| 14853 | TGTA | T | Deletion | V472 deletion | RdRP | 1 | 141 | No |
| 14877 | C | T | Synonymous | Y479 | RdRP | 1 | 124 | Yes, Gámbaro *et al.* (2020) |
| 14889 | C | T | Synonymous | Y483 | RdRP | 1 | 117 | No |
| 15738 | C | T | Synonymous | F766 | RdRP | 1 | 354 | No |
| 16716 | C | T | Synonymous | D160 | helicase | 1 | 93 | No |
| 23403 | A | G | Non-synonymous | D614G | spike (S) | 6 | 541 | Yes, Numerous |
| 25397 | AT | A | Deletion | Frameshift | ORF3 | 1 | 243 | No |
| 26753 | C | T | Synonymous | T77 | membrane (M) | 1 | 43 | No |
| 27190 | A | G | Synonymous | Silent, at stop codon | membrane (M) | 1 | 96 | No |
| 27317 | A | G | Non-synonymous | N39S | ORF6 | 1 | 101 | No |
| 27345 | A | G | Synonymous | K48 | ORF6 | 1 | 101 | No |
| 27392 | A | G | NONE | NONE | non-coding region between | 1 | 100 | No |
|  |  |  |  |  | ORF6 and<br>ORF7a |  |  |  |
| 27394 | A | G | Synonymous? | Alternative start<br>codon? (AUG --><br>GUG) | ORF7a | 1 | 94 | No |
| 27398 | A | G | Non-synonymous | K2R | ORF7a | 1 | 99 | No |
| 27400 | A | G | Non-synonymous | I3V | ORF7a | 1 | 96 | No |
| 27487 | A | G | Non-synonymous | K32E | ORF7a | 1 | 95 | No |
| 28242 | G | A | Non-synonymous | V117I | ORF8 | 2 | 476 | No |
| 28250 | T | TCTG | Insertion | 120/121 FI --> LFS<br>+ loss of stop<br>codon | ORF8 | 1 | 143 | Might be similar to<br>reported mutations by<br>Zinzula (2021) |
| 28253 | CA | C | Deletion |  |  | 1 | 139 |  |
| 28311 | C | T | Non-synonymous | P13L | nucleocapsid<br>(N) | 2 | 450 | Yes, Tablizo *et al.* (2020) |
| 28881 | GGG | AAC | Non-synonymous | 203/204 RG --> KR | nucleocapsid<br>(N) | 2 | 88 | Yes, Tablizo *et al.* (2021) |
| 29421 | C | T | Non-synonymous | P383L | nucleocapsid<br>(N) | 1 | 51 | Yes, several |

## DISCUSSION

Wastewater-based surveillance of SARS-CoV-2 has been proven to be a useful surveillance tool for COVID-19 (Daughton, 2020). In addition, WBE can support clinical surveillance by detecting pre-symptomatic and asymptomatic COVID-19 cases (Wannigama et al., 2021). It is also an additional indicator for COVID-19 prevalence trends in the sampled neighborhood or community (Peccia et al., 2020).

In this study, SARS-CoV-2 was detected in the wastewaters of *barangays* from Davao City, Philippines, regardless of the COVID-19 transmission risk category of the *barangays* or presence or absence of reported cases. The weekly *barangay-level* COVID-19 transmission risk classification released by the City Government of Davao City is based on ADAR and 2WGR. Weekly attack rates from October to December 2020 in the involved barangays in this study correlated with SARS-CoV-2 Ct values. However, in our study, no striking temporal pattern was observed. Furthermore, WBE detected SARS-CoV-2 RNA in barangays even when the attack rate was low (one new case per 10,000 persons). SARS-CoV-2 RNA was also detected in two of the most densely populated barangays included in this study, even when no new cases were reported for the corresponding week. Wannigama et al. (2021) reported a similar result in Thailand, where SARS-CoV-2 genome copies were detected in wastewater from Bangkok’s City Center and neighboring suburbs even when cumulative cases were zero from July 2020 to November 2020. Trottier et al. (2020) also did not observe a temporal correlation between the SARS-CoV-2 genome copies in wastewater and the COVID-19 incidence in Montpellier, France, as high levels of average SARS-CoV-2 genome copies were detected weeks before the COVID-19 cases increased in July 2020. This finding is possible because of fecal shedding in asymptomatic and pre-symptomatic COVID-19 cases, the variable amount of shed SARS-CoV-2 among infected persons (Trottier et al. 2020; Wannigama et al. 2021), or differences in testing coverage in the community. Since all active cases of COVID-19 in Davao City are immediately taken to one of the eleven Temporary Treatment and Monitoring Facilities (TTMFs) shortly after testing positive via RT-PCR (City Government of Davao, 2020), we hypothesize that the SARS-CoV-2 RNA detected in the community wastewater may come from either pre-symptomatic, or asymptomatic cases, or symptomatic individuals who did not self-report to their local health monitoring unit.

Over 70% of confirmed COVID-19 active cases in Davao City by the end of 2020 were asymptomatic cases (DOH Davao Region, 2021). This can be attributable to aggressive active-case finding, intensified contact tracing, and free community testing via RT-PCR in the City, whereby even asymptomatic contacts of confirmed COVID-19 cases are tested and required to self-isolate (City Government of Davao, 2021). In cities or municipalities where clinical testing capacity and healthcare resources are low, both symptomatic and asymptomatic COVID-19 infections may be missed by clinical diagnostics (testing via RT-PCR). Even without testing resource limitations, COVID-19 infections in patients who purposely do not report having symptoms, and hence, do not get tested for COVID-19 because of fear of social stigma (Michael-Kordatou et al., 2020), or isolation and quarantine (Larsen and Wigginton, 2020) will not be captured by clinical surveillance and contact tracing. Wastewater-based epidemiology offers a complementary public health surveillance method that provides community infection data faster using less resources than individual RT-PCR testing (Daughton, 2020). With long-term periodic community wastewater testing for SARS-CoV-2, WBE can present an unbiased snapshot of COVID-19 cases at the community level, whether asymptomatic, pre-symptomatic, or symptomatic (Wu et al., 2020). WBE can also be useful in suburban and rural communities, especially in low-resource settings, where people may have limited access to healthcare (Larsen and Wiggington, 2020).

Genomic surveillance in wastewater was attempted in this pilot work, and an average read depth of genome coverage was calculated to be ∼20x, which indicates that SARS-CoV-2 genomes were indeed sequenced reasonably and accurately (Goldman and Domschke, 2014). However, fragmented genomes were generated probably due to 1) the problematic nature of the sample, which may have been composed of degraded viral genetic material, and 2) the high Ct values (above 30) of the samples, which is known to be technically challenging for whole genome sequencing (Crits-Christoph *et al*., 2020). Nevertheless, we analyzed the sequence data as an exploratory frame of reference for future investigations. Furthermore, to ensure high-quality analysis despite the fragmented sequences, the threshold for variant calling was performed with a total of 30x coverage (Izquierdo-Lara *et al*., 2021).

Our wastewater surveillance in Davao City detected SARS-CoV-2 non-synonymous mutations which have been previously reported in the Philippines: [1] the nsp3 T2016K, RdRp A97V, and nucleoprotein P13L, which were first detected in March 2020 (Tablizo *et al*., 2020); [2] the nucleoprotein R203K and G204R, which were first detected in January 2021 (Tablizo *et al*., 2021); and [3] the spike D614G which has become globally dominant early in the pandemic (Korber *et al*., 2020). In addition, other mutations reported in other countries (nsp2 P129S, RdRp A449V, and 120/121FI→ LFS) (Sengupta *et al*., 2020, Wang *et al*., 2020, Zinzula e*t al*., 2021) were also detected in this study.

We also detected mutations that were not yet reported by clinical surveillance. One of these mutations is the V117I non-synonymous mutation at ORF8 represented in two samples (total of 476 observations). ORF8 is an accessory protein for viral capsid formation and is also implicated in the host immune response towards SARS-CoV-2 (Santerre *et al*., 2020; Zinzula, 2021). Another mutation of interest is the deletion at position 11083 located in the nsp6 gene region which is represented in two samples (total of 605 observations). Previous publications in the Philippines and elsewhere (Tablizo *et al*., 2020; Aiewsakun *et al*., 2020; Chaw *et al*., 2020) have reported a non-synonymous mutation at this position, but not the observed deletion in this study. The viral protein encoded by nsp6 assists in the formation of vesicles for viral replication (Santerre *et al*., 2020). Other previously unreported non-synonymous mutations and indels were detected in only one site, such as for the vesicle-forming nsp3 and nsp6, the polymerase RdRp, and the accessory proteins ORF3, ORF6, and ORF7a.

Except for the D614G mutation, there was a notable absence of mutations within the spike gene, previously noted for VOCs and the Philippine P3 variant. Unfortunately, most of the regions covered by these mutations were not successfully sequenced, thereby preventing any conclusion from being made on these mutations. On the other hand, the sub-regions for S94F, 141-143del, P681H/R, K417N/T, E1092K, and H1101Y were covered in the sequence data, and the corresponding mutations were absent.

Altogether, our genomic surveillance in wastewater confirmed the presence of previously reported mutations while identifying mutations not yet registered in clinical surveillance, albeit the current strategy has large room for improvement considering the fragmented nature of the sequence data. Although the Philippines has a national genomic biosurveillance program, only representative clinical samples are included (DOH, 2021), thereby introducing the possibility of excluding other relevant circulating SARS-CoV-2 variants. Wastewater genomic surveillance can complement clinical surveillance by providing additional genomic information about circulating and emerging mutations using lesser resources, given that pools of individuals are represented in every wastewater sample used for WGS.

This pilot application identified a unique set of challenges for wastewater surveillance in the Philippines. COVID-19 WBE has been reported in over 50 countries worldwide, and 94% of these countries belong to the high and upper-middle income countries that have centralized wastewater treatment plants (WWTPs) (Naughton et al., 2021 PREPRINT). With WWTPs, accurate data about the population served by the specific treatment plant is available, and biological, physicochemical, and hydrologic data are regularly monitored, permitting near-real-time spatio-temporal trend analysis on COVID-19 transmission in the served population (Michael-Kordatou, Karaolia, and Fatta-Kassinos, 2020). Unlike high and upper-middle income countries, most cities in the Philippines, including Davao City, do not have centralized wastewater treatment facilities. Over 90% of households in Davao City are connected to private or communal septic systems (JICA, 2018), and sewer pipes carry primary-treated wastewaters along with surface runoff onto the draining natural bodies of water. One of the challenges in this study was obtaining reliable estimates of the contributing population to the wastewaters collected since data on sewer pipe networks in the *barangays* were fragmented, and only personal knowledge of barangay personnel about sewer networks was available as of writing. Cities without centralized WWTP that plan to implement WBE, whether for COVID-19 or other water-related infectious diseases, must ensure that a comprehensive map of sewer pipelines is first generated to identify the population contributing to the wastewater accurately. However, WBE can also be used for non-intrusive targeted site testing for COVID-19 infections. For example, wastewater samples from residential buildings were tested for SARS-CoV-2 in Singapore (Wong et al., 2021) and Hong Kong (Sharon, 2021). Positive test results prompted their respective health agencies to implement compulsory testing of residents and isolation of infected persons, thereby limiting the spread of COVID-19 in the tested community.

The location of wastewater sampling was also crucial in this study. Sample collection at sites close to the Davao Gulf (i.e., 76-A Bucana, Leon Garcia, Monteverde, and 23-C) was greatly influenced by tidal changes. During high tide, the sewer pipes become partially submerged in seawater (or riverine water at 76-A Bucana), and the direction of wastewater flow is inward, which reverses during low tide. Besides dilution, saltwater intrusion to wastewater may hasten the decay of enveloped viruses like SARS-CoV-2 (Pinon and Vialette, 2018) and may affect the levels of detectable viral RNA in the samples. Hence, sample collection was done only during low tide to reduce wastewater dilution with seawater. Therefore, when planning WBE using sewer lines that open to a river or the sea, sewer pipelines that are consistently above the level of a river or seawater are ideal.

Furthermore, anthropogenic factors such as industrial discharge and livestock wastes have been shown to alter the water quality and may influence nutrient loads, coliforms, and chemical composition (Khatri and Tyagi, 2015). O’Brien and colleagues (2017) also found that the abundance and diversity of viruses in wastewater varied with their environment. We are currently evaluating physico-chemical, hydrologic, and anthropogenic factors’ contributions to recovering SARS-CoV-2 RNA from community wastewater in Davao City.

Another factor to consider in WBE is the sampling strategy. In most cities, the sewage flow rate is greatest during the morning and the evening, and a 24-hour composite sampling can catch these peak flows with the most reliable mean viral levels in the wastewater (Michael-Kordatou et al., 2020). However, composite samples may also result in reduced viral titers compared to grab sampling (Michael-Kordatou et al., 2020, Nemudryi et al., 2020 PREPRINT) because of the dilution of viruses (Napit et al., 2021 PREPRINT). In addition, composite sampling is also logistically challenging, especially without expensive automated wastewater collectors. In this study, grab samples were collected between 8 AM to 3 PM, depending on the site’s distance from the laboratory and the changing of tides (for sites near the Davao Gulf). For future studies, the collection of grab samples during peak hours of human toilet use (e.g., 7 AM to 9 AM) is an alternative method that can improve the chances of detecting fecally shed SARS-CoV-2, especially in low-incidence and low-resource areas (Napit et al., 2021 PREPRINT).

Another critical step in WBE is choosing a concentration method for viruses (Michael-Kordatou et al., 2020). Wastewater is an environmental matrix with high chemical and biological complexity that may affect viral recovery (Shi, Pasco, and Tarabara, 2017). Since most fecally-shed SARS-CoV-2 viruses are unprotected or non-intact forms (Wurtzer et al., 2020), they are more susceptible to degradation. The method for the concentration of viruses from wastewater used in this study is a combination of PES filtration and PEG-NaCl precipitation, modified from Wu et al. (2020). However, RNA recovery was not computed in this study; RNA levels of wastewater-concentrated viruses correlated with the SARS-CoV-2 Ct values. Ahmed et al. (2020) reported that using PEG precipitation can yield RNA recoveries ranging from 26.7 to 65.7% using a SARS-CoV-2 surrogate virus (murine hepatitis virus). RNA recovery is an essential measure of how much RNA is available for downstream SARS-CoV-2 detection (CDC, 2020), and this may depend on several factors, such as the volume of wastewater samples collected, sample processing (including viral concentration), and RNA extraction method chosen (Kantor et al., 2021). Michael-Kordatou et al. (2020) further recommended processing higher volumes of wastewater in low COVID-19 incidence areas to increase the chances of SARS-CoV-2 detection.

The method used for detection has to be considered. The standard markers used for SARS-CoV-2 detection in wastewater are the N1, N2, and N3 genes and E Sarbecovirus gene (Kitajima et al. 2020). Our pilot investigation suggests using either the N or RdRp gene, as they had consistently higher rates of detection (75-100%) relative to E (62.5%) both in the community and hospital wastewaters. Ct values in this study were also within the previously reported range of 27 to almost 40 (Ahmed et al., 2020a and 2020b, Bar-Or et al., 2020, Kocamemi et al., 2020, Kumar et al., 2020, Napit et al., 2021, Wu et al., 2020). Because of the absence of costly quantification standards for RT-PCR, quantification of viral genome copies per volume of wastewater and, consequently, the determination of analytical specificity (i.e., limit of detection, LOD) were not performed in this pilot study. Although the AllPlex™ 2019-nCOV assay (Seegene Inc., 2020) was developed for clinical applications using respiratory samples, we demonstrate its applicability for SARS-CoV-2 detection in feces-contaminated environmental samples, as has been done in Nepal (Napit et al. 2021 PREPRINT). Subsequent WBE studies must adhere to the Environmental Microbiology Minimum Information (EMMI) for publication of Quantitative Real-time PCR Experiments which describes essential and desired qPCR information, such as RNA yield, contamination assessment, inhibition testing, LOD determination, and repeatability, that researchers must report to ensure the validity and reliability of qPCR studies (Borchardt et al., 2021).

It is unclear if the wastewater samples that were negative for SARS-CoV-2 genes in this study collected on weeks when both new and active cases were reported were indeed true negatives. In addition, more robust WBE data would have been reported if the determination of RNA recovery and LOD and PCR inhibition studies were performed and correlated with the detection of SARS-CoV-2 target genes. Nonetheless, to our knowledge, this is the first paper to report COVID-19 wastewater surveillance in the Philippines and the Western Pacific region.

## CONCLUSION AND RECOMMENDATIONS

In summary, SARS-CoV-2 RNA was detected in community-level wastewater from the six barangays of Davao City, even when the barangays were classified as having a Low Risk of COVID-19 transmission and no new cases were reported. In addition, previously reported mutations and novel non-synonymous mutations were also detected from whole-genome sequencing of wastewater. These results highlight the potential of systematic wastewater-based epidemiology for SARS-CoV-2 as a complementary approach to clinical surveillance even in a low-sanitation metropolitan like Davao City, Philippines. Viral load per community must be estimated to generate COVID-19 transmission trends and assist the local government in planning and evaluating the appropriate public health interventions. Future studies must also explore the effect of different physico-chemical, biological, hydrologic, and anthropogenic parameters on viral detection and quantification following the EMMI guidelines. When planning to adopt WBE for COVID-19 in areas without WWTPs and with low testing resources, cities and/or research groups must consider: 1) knowledge or a map of sewer lines and available population data contributing to the wastewater to be monitored, 2) proper site selection and appropriate sampling design considering the local conditions, 3) the impact of onsite anthropogenic and environmental factors on detection, 4) optimized methods for virus concentration and detection, and 5) adequate resources for field and laboratory work.

## Data Availability

Raw data available in the supplemental materials

## ACKNOWLEDGEMENTS

This project was supported by the University of the Philippines Mindanao In-House Research Grant, the DOST -Philippine Council for Health Research and Development through the Philippine Genome Center Mindanao, the Just One Giant Lab through the Project Accessible Genomics, New England BioLabs Inc., and USAID PEER subaward number (2000009924).

The authors would also like to thank the Davao City Health Office, the local government units under the City Government of Davao, and partner hospitals for their support and assistance, and Dr. Diana Aga for discussions on sample collection and analyses. The authors would also like to acknowledge the following for their expert advice on nanopore and wastewater sequencing: Ralf Dagdag and Matthew Redlinger from the Bortz Virology Laboratory at the University of Alaska Anchorage; Dr. Amanda Warr from the Roslin Institute of the University of Edinburgh; Dr. Nicole Wheeler from the University of Birmingham; Dr. Lara Urban, co-founder of *PuntSeq*; and Dr. Joe Russell from MRIGlobal.

