## Supplemental Tables for "Wastewater-Based Epidemiology and Whole-Genome Sequencing for Community-Level Surveillance of SARS-CoV-2 in Selected Urban Communities of Davao City, Philippines: A Pilot Study"

**Supplemental Material**

Table S1. Sampling, extraction, and SARS-CoV-2 detection data of wastewater samples from Davao City, Philippines.

| Barangay | Date of collection | Time of collection | RNA Concentra-  tion (ng/uL) | CT values | | | Interpreta-tion | Sequencing (Y/N) |
| --- | --- | --- | --- | --- | --- | --- | --- | --- |
|  |  |  |  | E | N | RdRP |  |  |
| 23-C | 11/13/20 | 2PM | 28.0 | 31.793 | 32.154 | 34.636 | POSITIVE | Y |
|  | 11/27/20 | 10AM | 7.8 | UND | UND | UND | NEGATIVE | N |
|  | 12/4/20 | 12PM | 23.2 | UND | 38.739 | 35.283 | POSITIVE | N |
|  | 12/11/20 | 11AM | 35.8 | UND | UND | 38.597 | POSITIVE | N |
| Leon Garcia | 11/13/20 | 11AM | 27.2 | 31.69 | 31.257 | 33.484 | POSITIVE | Y |
|  | 11/27/20 | 9AM | 19.4 | 37.088 | 35.591 | 35.008 | POSITIVE | N |
|  | 12/4/20 | 11AM | 40.2 | 39.727 | 31.702 | 31.893 | POSITIVE | N |
|  | 12/11/20 | 10AM | 36.6 | 32.037 | 29.406 | 31.264 | POSITIVE | Y |
| Monteverde | 11/13/20 | 11AM | 31.0 | 32.13 | 32.375 | 35.619 | POSITIVE | Y |
|  | 11/27/20 | 8AM | 22.8 | 31.031 | 31.909 | 33.814 | POSITIVE | Y |
|  | 12/4/20 | 10AM | 40.0 | 32.879 | 30.354 | 32.941 | POSITIVE | Y |
|  | 12/11/20 | 9AM | 37.4 | 37.709 | 37.414 | 38.888 | POSITIVE | N |
| 76-A Bucana | 11/9/20 | 8AM | 16.4 | 36.636 | 34.089 | 36.538 | POSITIVE | N |
|  | 11/23/20 | 8AM | 10.6 | 39.425 | 35.362 | 33.87 | POSITIVE | N |
|  | 11/30/20 | 9AM | 10.2 | 39.425 | 35.362 | 33.87 | POSITIVE | N |
|  | 12/7/20 | 2PM | 24.0 | UND | 38.266 | UND | POSITIVE | N |
| Matina Crossing | 11/9/20 | 11AM | 32.6 | 29.945 | 30.98 | 32.92 | POSITIVE | Y |
|  | 11/23/20 | 11AM | 10.6 | UND | 35.771 | 32.572 | POSITIVE | N |
|  | 11/30/20 | 11AM | 22.8 | 35.55 | 32.002 | 32.19 | POSITIVE | Y |
|  | 12/7/20 | 12PM | 21.2 | 33.09 | 34.124 | 33.225 | POSITIVE | N |
| Mintal | 11/9/20 | 1PM | 22.6 | UND | 36.429 | 37.261 | POSITIVE | N |
|  | 11/23/20 | 1PM | 10.6 | UND | UND | UND | NEGATIVE | N |
|  | 11/30/20 | 1PM | 11.8 | UND | UND | 34.005 | POSITIVE | N |
|  | 12/7/20 | 3PM | 10.2 | UND | 35.457 | 35.997 | POSITIVE | N |
| Hospital 1 | 11/9/20 | 11AM | 18.0 | 33.951 | 34.845 | UND | POSITIVE | N |
|  | 11/23/20 | 9AM | 11.2 | 35.463 | 34.842 | 36.103 | POSITIVE | N |
|  | 11/30/20 | 10AM | 22.4 | 35.376 | 30.949 | 31.670 | POSITIVE | N |
|  | 12/7/20 | 10AM | 16.4 | 33.032 | 32.591 | 31.703 | POSITIVE | Y |
| Hospital 2 | 11/13/20 | 9AM | 13.6 | 36.391 | 35.989 | 39.009 | POSITIVE | N |
|  | 11/27/20 | 11AM | 12.4 | UND | 36.893 | UND | POSITIVE | N |
|  | 12/4/20 | 8AM | 11.0 | UND | 37.475 | 34.026 | POSITIVE | N |
|  | 12/11/20 | 10AM | 16.4 | 37.64 | 36.362 | 33.138 | POSITIVE | Y |

Legend: UND, undetermined, Ct greater than 40.

Table S2. Regression analysis of N gene Ct prediction based on RNA concentration and Attack Rate.

|  | *Coefficients* | *95% CI* | *Standard Error* | *P-value* |
| --- | --- | --- | --- | --- |
| Intercept | 43.857 | [39.450,48.264] | 2.119 | 0.0000 |
| RNA concentration | -0.201 | [-0.355,-0.047] | 0.074 | 0.0129 |
| Attack Rate | -7.160 | [-11.856,-2.464] | 2.258 | 0.0046 |

Table S3. Regression analysis of E gene Ct prediction based on RNA concentration and Attack Rate.

|  | *Coefficients* | *95% CI* | *Standard Error* | *P-value* |
| --- | --- | --- | --- | --- |
| Intercept | 49.037 | [44.401, 53.675] | 2.229 | 0.0000 |
| RNA concentration | -0.256 | [-0.418, 0.094] | 0.0779 | 0.0035 |
| Attack Rate | -9.564 | [-14.505, -4.623] | 2.3758 | 0.0006 |

Table S4. Regression analysis of RdRP gene Ct prediction based on RNA concentration and Attack Rate.

|  | *Coefficients* | *95% CI* | *Standard Error* | *P-value* |
| --- | --- | --- | --- | --- |
| Intercept | 40.468 | [36.140, 44.800] | 2.082 | 0.0000 |
| RNA concentration | -0.101 | [-0.252, 0.050] | 0.072 | 0.1796 |
| Attack Rate | -4.902 | [-9.520, -0.290] | 2.218 | 0.0384 |

Table S5. Summary of SARS-CoV-2 genome assembly statistics and corresponding CoV-GLUE analyses.

| Sample (location and sampling date) | Read depth of genome coverage | Percent genome fraction coverage | Lineage assignment (CoV-GLUE) | Total likelihood weight ratio (lineage assignment) |
| --- | --- | --- | --- | --- |
| 76-A (11-23-20) | 20.74 | 17.08 | B.1.p11 | 63.5981 |
| Matina Crossing (11-09-20) | 69.18 | 50.06 | B.6 | 100.00 |
| Matina Crossing (11-30-20) | 0.43 | 1.16 | B.1 | 59.375 |
| Leon Garcia (11-13-20) | 18.37 | 19.34 | B.1 | 95.3125 |
| Leon Garcia (12-11-20) | 15.23 | 13.61 | B.1 | 95.3125 |
| Monteverde (11-13-20) | 20.83 | 10.34 | B.1.1 | 95.4147 |
| Monteverde (11-27-20) | 29.34 | 20.37 | B.1.1 | 95.3886 |
| Monteverde (12-04-20) | 20.54 | 16.63 | B.1 | 95.3125 |
| 23-C (11-13-20) | 15.66 | 9.00 | B.1 | 73.4375 |
| Hospital 1 (12-07-20) | 1.65 | 2.32 | B.1 | 68.75 |
| Hospital 2 (12-11-20) | 7.16 | 4.56 | B.1 | 65.625 |
| Negative control | 0 | 0 | - | - |

Table S6. Information on the detection of notable spike gene mutations found in Variants of Concern (VOCs) and the Philippine P.3 variant within wastewater-detected SARS-CoV-2 from Davao City, Philippines on Nov- Dec 2020.

| **Notable mutations of the spike gene** | **Attributed Effect** | **Covered in the sequence data?** | **Mutation present in the sequence data?** | **Reference(s)** |
| --- | --- | --- | --- | --- |
| Global VOCs |  |  |  |  |
| K417N/T | Unclear (probable immune evasion) | Yes (2 samples) | No | Winger and Caspari, 2021 |
| L452R | Immune evasion | No | n/a |  |
| E484K/Q | Immune evasion | No | n/a |  |
| N501Y | Increased transmissibility | No | n/a |  |
| D614G | Increased transmissibility | Yes (6 samples) | Yes |  |
| P681H/R | Unclear (probable increased transmissibility) | Yes (4 samples) | No |  |
| V1176F | Unclear (probable increased disease severity) | No | n/a | Farkas *et al*., 2020 |
| Philippine P.3 variant |  |  |  |  |
| S94F | Unknown | Yes (2 samples) | No | Tablizo *et al.*, 2021; Bascos *et al.*, 2021 |
| 141-143del | Unclear (probable increased stability to spike monomers) | Yes (2 samples) | No |  |
| 243-244del | Unknown | No | n/a |  |
| E1092K | Unclear (probable increased stability to spike trimers) | Yes (2 samples) | No |  |
| H1101Y | Unclear (probable increased stability to spike monomers) | Yes (2 samples) | No |  |
